# Cohort profile: Medical Birth Registry at Kilimanjaro Christian Medical Centre (KCMC), Tanzania 2000-2023

**DOI:** 10.1101/2025.01.09.25320304

**Authors:** Daire Buckley, Ali Khashan, Innocent Mboya, Modesta Mitao, Gustav Nkya, Allen Senkoro, Allen Lyimo, Francis M. Pima, Lucy Munishi, Simon Woodworth, Raheli Manongi, Gileard Masenga, Michael Mahande, Clifford Silver Tarimo, Anne Kjersti Daltveit, Rolve Terje Lie, Bariki Mchome, Blandina T. Mmbaga

## Abstract

**Purpose:** Birth registry data is integral to healthcare providers, and policymakers. The primary aim of the data is to record, track, and monitor maternal and new-born outcomes. The objectives of the Kilimanjaro Christian Medical Centre (KCMC) medical birth register are to serve as tools for monitoring births and key obstetric outcomes such as caesarean sections, and maternal and perinatal deaths for clinical, administrative and research purposes. (1) Subsequent births and births can also be followed to serve to bridge a gap in maternal and new-born health information within the Kilimanjaro Region of Tanzania.

**Participants:** Participants are women who give birth at the Obstetrics and Gynaecology Department of The Kilimanjaro Christian Medical Centre (KCMC) in the Moshi Municipality, Kilimanjaro Region, Northern Tanzania.

**Methods:** The KCMC Medical Birth Registry was established in 2000. Initially, it was based on a desktop database. In 2020 it was transformed into a digital system, Castor EDC platform, and in January 2024 it was moved to the open-source DHIS2 system. There were over 60,000 births from 2000-2019 were recorded. A total of 8,042 births were recruited from the KCMC facility from January 2020 to December 2023. Participants were interviewed by trained research midwives within 24 hours of giving birth. A standardised questionnaire is used during interviews to capture relevant information on maternal and paternal background characteristics, maternal information before and during the current pregnancy, and obstetric/ clinical information about births (including singletons and multiple births).

**Findings to date:** Of the total cohort, 4002 (49.8%) participants had a spontaneous vaginal delivery, while almost a third of participants delivered by caesarean section. Instrumental assistance was primarily in the form of a forceps delivery (11.9%), followed by ventouse delivery (1.3%). Additional neonatal outcomes included child status (live/stillborn), and the Apgar score among a long list of variables. A varied proportion of data was missing (ranging from 3% to 7%) from both clinical and postpartum variables. Since 2000, more than 40 peer-reviewed papers have been published using this database.

**Future plans:** As the methods of the birth registry data collection continue to improve, the data that is collected from the birth registry systems will aid policymakers in monitoring the country’s progression and develop targets for strengthening healthcare systems at both a national and international level. From a research perspective, the data is available for international collaboration for conducting high-quality research into the cause and prevention of adverse pregnancy outcomes in low-resource settings.

## Introduction

The Kilimanjaro Christian Medical Centre (KCMC) medical birth registry was established in 2000. It acts as a database for collecting comprehensive perinatal information about births that occur at the hospital. The registry is part of a larger effort to monitor obstetric outcomes in Tanzania (1). Birth data play a key role in monitoring trends and patterns of key health indicators in maternal and child health, and the identification of risk factors associated with adverse outcomes including maternal mortality rates, infant mortality rates, and fertility rates. Low- and lower-income countries carry the greatest global burden of maternal and perinatal mortality, and it is estimated that 70% of all global maternal deaths in 2020 occurred in sub-Saharan Africa. According to the World Health Organisation (WHO), this equates to 545 maternal deaths per 100,000 live births. Since there is a reliance on paper-based records in health settings in many low-income countries, it is likely a knowledge gap exists in relation to the true burden of MMRs due to inaccessible documentation of vital statistics. Between 2000 and 2020 the WHO reported the overall proportion of maternal mortality rates (MMR) had stalled in 133 countries and there was an increase in MMR in 17 countries (2) .While over those two decades, MMRs declined by 69.8%, in comparison to other countries, the maternal mortality rates remain high estimated at 238 deaths per 100,000 live births in Tanzania (WHO., 2023). According to a study involving 34 public hospitals in Tanzania conducted between July and December of 2016, there was a noticeable surge in hospital MMR in public hospitals of Tanzania within a ten-year time frame (2006 to 2015). The deaths accounted for approximately 5% of all maternal in-hospital mortalities within the child-bearing age group. Direct causes of maternal mortality were recorded as eclampsia, obstetric haemorrhage, and maternal sepsis, while the highest percentage of indirect causes of maternal deaths were anaemia and cardiovascular diseases (3) .

Therefore, these data capture and collection methods using electronic health records provided accessible and effective coverage of delivery outcome patterns and trends within obstetric services and identified the predominant risk factors for expectant mothers and highlighted to healthcare providers at a local level in a low-income setting. From this perspective, the KCMC Medical Birth Registry (KCMC MBR) becomes a vital tool for monitoring these trends and plays an important role in enhancing obstetric care.

Thus far, the implementation of the KCMC MBR has demonstrated the need for accurate and full medical records in a limited resource setting and the collected data has contributed a multitude of research publications.

The aim of this cohort profile is threefold, firstly, to provide an updated and comprehensive overview and description of the KCMC Medical Birth Registry in Tanzania, outlining data collection and follow-up procedures; secondly to present current findings derived from the MBR and thirdly, to provide details on accessing the registry data for research purposes. This detailed profile aims to enhance transparency and understanding of the KCMC MBR operations but also facilitate collaboration and utilisation of robust data for further research within the scientific community.

## Methods

### Cohort description, location, and key dates

KCMC is located in the Moshi Municipality, Kilimanjaro Region, Northern Tanzania, and is one of four zonal referral hospitals in Tanzania. The 2022 Census Results recorded the population of Tanzania at 61,741,120. (4) The hospital serves as a referral hospital for seven administrative regions in North Tanzania, serving a catchment population of over 11 million people. Births at KCMC were managed by paper-based health information system until 2019. The KCMC Birth Registry was initiated as a pilot project in collaboration with The Medical Birth Registry of Norway, University of Bergen, and the Kilimanjaro Christian Research Institute (KCRI). (5) Since 2000, perinatal data on maternal and infant outcomes has been recorded via Microsoft Access Data. Since 2020, the Castor EDC Platform has been used to record data and in January 2024, the MBR was moved to the open-source DHIS2 platform. The KCMC MBR has recorded an average 2000-2500 births per year since its establishment. Over 60,000 births have been recorded in the KCMC MBR. (6)

Births are mainly from the local population of the Moshi Municipality and Moshi Rural District, with 80% of the women who deliver in KCMC being self-referred, and the surrounding districts admit the remaining referrals for management of obstetric complications. (7,8); The KCMC MBR was established to monitor perinatal health and the quality of care delivered at KCMC; however, the collected data is also a vital source of information on maternal and infant outcomes for babies born at the facility every year for healthcare providers, funders and policymakers.

### Eligibility criteria and data collection

The KCMC MBR eligibility is open to all mothers who gave birth in the Obstetrics and Gynaecology Department of KCMC and consented for their data to be included in the MBR. Interviews with each consenting mother are conducted by specially trained research midwives, within the first 24 hours after delivery. Where women have undergone a delivery complication, interviews are conducted on the second or third day after birth, depending on their condition. A standardised questionnaire (see appendix B) is used during interviews to capture relevant information on the mothers referral to the hospital (be it self-referral/transferral under medical advice for specialist care), and factors pertaining to maternal and paternal socio-demographic information such as education attainment, occupation, health status, smoking and alcohol consumption, HIV/Syphilis status (if known), and maternal reproductive history (prime/multiparous; births, miscarriage(s), ectopic pregnancy, and outcomes), are collected.(For full access to the variable list see Appendix A.) Only mothers who consented to participate are included in the MBR. Data on the mode of delivery (caesarean or vaginal), as well as complications, interventions, and outcomes are collected. In addition, data on the child status (live or stillborn), birth weight, height, and Apgar score of the offspring is also recorded. Additional data is abstracted from the antenatal care cards (ANC) and hospital records of the mother. All women are assigned a unique identification number which can be used to link mother and child information and monitor the reproductive life of the woman, including subsequent births, or additional adverse pregnancy outcomes if any. (5, 6)

## Results

### Baseline characteristics of study participants

Baseline characteristics of 8,042 study participants that were collected from January 2020 to December 2023 and are summarised in Table 1. Briefly, most participants (71.4%) were categorised between the ages of 20-34 years. Almost half of the participants (47%) resided in an urban area, while 37.9% resided rurally, and 12.9% resided in semi-rural areas. Most women were employed, married, and had attended a form of education. The predominant tribe was the Chagga tribe (50.3%). 42.2% of participants identified as protestant. Close to 100% of the cohort did not smoke, while 82.9% did not consume alcohol before pregnancy.

**Table 1:** Baseline Socio-demographic characteristics of study participants who delivered at KCMC 2020-2023 (N=8,042)

| Variable | n | % |
| --- | --- | --- |
| **Maternal age (n=7588)** |  |  |
| < 20 years | 288 | 3.6 |
| 20-34 years | 5764 | 71.4 |
| ≥ 35 years | 1536 | 19.1 |
| Missing | 454 | 5.9 |
| **Area of residence (n=7878)** |  |  |
| Rural | 3054 | 37.9 |
| Urban | 3782 | 47 |
| Semi-urban | 1042 | 12.9 |
| Missing | 164 | 2.2 |
| **Education level (n=7868)** |  |  |
| None | 75 | 1 |
| Primary | 1695 | 21.1 |
| Secondary | 3471 | 43.1 |
| Higher | 2627 | 32.6 |
| Missing | 174 | 2.2 |
| **Employment status (n=7878)** |  |  |
| Housewife | 876 | 10.9 |
| Farmer | 548 | 6.8 |
| Service | 535 | 6.7 |
| Business | 2794 | 34.7 |
| Professional | 2715 | 33.7 |
| Student | 395 | 4.9 |
| Others | 15 | 0.2 |
| Missing | 164 | 2.1 |
| **Relationship status (n=7877)** |  |  |
| Married | 6335 | 78.8 |
| Single | 1491 | 18.5 |
| Widowed | 33 | 0.4 |
| Remarried | 7 | 0.1 |
| Divorced | 5 | 0.1 |
| Polygamous | 6 | 0.1 |
| Missing | 165 | 2.0 |
| **Mother's tribe (n=7874)** |  |  |
| Chagga | 4042 | 50.3 |
| Pare | 921 | 11.5 |
| Masai | 218 | 2.7 |
| Other | 2693 | 33.5 |
| Missing | 168 | 2 |
| **Maternal religion (n=7859)** |  |  |
| Catholic | 2977 | 37 |
| Protestant | 3395 | 42.2 |
| Muslim | 1319 | 16.4 |
| Other | 168 | 2.3 |
| Missing | 183 | 2.1 |
| **Smoker during pregnancy (n=8042)** |  |  |
| Yes | 40 | 0.5 |
| No | 8002 | 99.5 |
| **Alcohol consumption pre-pregnancy (n=7851)** |  |  |
| Yes | 1187 | 14.7 |
| No | 6664 | 82.9 |
| Missing | 191 | 2.4 |

### Clinical characteristics of study participants

The clinical characteristics of study participants are outlined in Table 2. Of all births studied, a total of 1591 (19.8%) of the study participants were categorised as anaemic, with haemoglobin values measured at <11g/dl. 398 (4.9%) of participants were found to be positive for Human Immunodeficiency Virus status (HIV status). 137 (1.8%) of the participants had gestational diabetes, while 230 participants (2.9%) were classified as having pregnancy hypertension, and 158 (1.7%) had eclampsia. 161 participants (2%) had malaria during their pregnancy.

**Table 2:** Clinical characteristics of study participants who delivered in KCMC (N=8042)

| Variable | n | % |
| --- | --- | --- |
| **Anaemic (g/dl) (n=7735)** |  |  |
| Yes (<11) | 1591 | 19.8 |
| No (≥11) | 6144 | 76.4 |
| Missing | 307 | 3.9 |
| **HIV status (n=7803)** |  |  |
| Positive | 398 | 4.9 |
| Negative | 7405 | 92 |
| Missing | 239 | 3.1 |
| **Gestational diabetes (n=7766)** |  |  |
| Yes | 137 | 1.6 |
| No | 7629 | 94.9 |
| Missing | 276 | 3.5 |
| **Pregnancy Hypertension (n=7752)** |  |  |
| Yes | 230 | 2.9 |
| No | 7522 | 93.5 |
| Missing | 290 | 3.6 |
| **Eclampsia (n=7752)** |  |  |
| Yes | 158 | 1.7 |
| No | 7594 | 94.4 |
| Missing | 290 | 3.6 |
| **Malaria in pregnancy (n=7774)** |  |  |
| Yes | 161 | 2 |
| No | 7613 | 94.7 |
| Missing | 268 | 3.3 |
| Abbreviations: Haemoglobin values <11g/dl are categorised as anaemic; HIV status: Human Immunodeficiency Virus status. |  |  |

Postpartum outcome data is outlined in Table 3. Of the total cohort, 4002 (49.8%) participants had a spontaneous vaginal delivery, while almost a third (29.2%) delivered by caesarean section. Interventions in the form of instrumental assistance were predominantly: forceps for 953 participants (11.9%), followed by ventouse delivery for 197 participants (1.3%). Neonatal outcomes of interest that were recorded include child status (live born/perinatal loss (referring to the death of an infant due to miscarriage, stillbirth, and neonatal death)), birth weight and gestational age of participants, and the Apgar score. A varied proportion of data was missing (ranging from 3% to 7%) from both clinical and postpartum variables.

**Table 3:** Postpartum outcomes of study participants in KCMC (N=8,042)

| Variable | n | % |
| --- | --- | --- |
| **Mode of delivery (n=7465)** |  |  |
| Spontaneous | 4002 | 49.8 |
| Vacuum | 107 | 1.3 |
| Forceps | 953 | 11.9 |
| Caesarean section | 2352 | 29.2 |
| Breech | 15 | 0.2 |
| Other | 36 | 0.5 |
| Missing | 577 | 7.7 |
| **Neonatal delivery status (n=7605)** |  |  |
| Live born | 6522 | 81.1 |
| Perinatal loss | 1075 | 14.1 |
| Missing | 445 | 4.8 |
| **Infant sex (n=7505)** |  |  |
| Male | 3814 | 47.4 |
| Female | 3655 | 45.5 |
| Unknown | 36 | 0.4 |
| Missing | 537 | 6.7 |
| **APGAR score (N=7457)** |  |  |
| 0-3 | 242 | 3 |
| 4-6 | 191 | 2.4 |
| 7-10 | 7024 | 87.6 |
| Missing | 585 | 7.3 |

## Discussion

The KCMC MBR provides valuable data source and insights into the delivery practices and maternal and neonatal outcomes at the KCMC healthcare facility. The MBR enables healthcare professionals to examine patterns in adverse pregnancy outcomes, identify risk factors associated with adverse obstetric outcomes and evaluate the impact of public health policies on these outcomes. The KCMC MBR provides a unique data source that enables the prediction of adverse outcomes to subsequently address the major concerns such as maternal and neonatal mortality rates within this region.

### Publications

Over 40 peer-reviewed publications have been published from data that had been collected from the KCMC Birth registry. Such data gives an insight into Tanzania’s reproductive health status by identifying the essential predictors of adverse maternal health issues like preterm birth, perinatal death, and adverse foetal risk factors such as the risk of stillbirth and low birth weight.

A recent publication used the KCMC birth registry data to measure the overall rate of recurrent preterm birth (24.4%) and identified pre-eclampsia, previous history of PTB, and prolonged inter-pregnancy intervals as statistically significant risk factors associated with recurrent pre-term birth. (9)

Another publication used the data to measure the impact of advanced maternal age (AMA) on perinatal outcomes. The study found that women categorised as AMA were more likely to deliver by caesarean section, were predisposed to have gestational diabetes, and were more likely to develop pre-eclampsia. (10)

The data has also enabled researchers to quantify the proportion of women in labour who were referred from district and regional hospitals (secondary-level health facilities) to a tertiary hospital for birth, resulting in almost half the population giving birth by caesarean section (45%). Timely referral to higher-level facilities were identified as a key component to preventing maternal death. The study also highlighted and called for improvements to be made at secondary-level facilities to better enable them to manage obstetric complications in emergencies. (11)

See Appendix C for full list of papers.

### Strengths and limitations

1. A limitation of this study is the missing data on study participants from the medical birth registry. Incomplete data may skew the results of studies, which could potentially affect the accuracy and completeness of the analysis and interpretation of findings. It is important to consider and address the missing data to ensure the reliability and validity of study results. Therefore, continued efforts are required to sustain and improve data collection and completeness on a long-term basis to mitigate the impact of missing data for future studies.
2. A strength of the registry is the unique identifier code given to women who participate, which enables researchers to monitor obstetric outcomes for women in subsequent births and compare the data to their previous births. This unique feature not only enhances data accuracy and follow-up but also enables a longitudinal analysis of trends and outcomes over time. This longitudinal tracking can offer valuable insights into maternal and infant health patterns, informing future interventions and policies aimed at improving overall outcomes in obstetrics and gynaecology. But also, we recommend the use of a national identification number for every woman so that we can link their important information such as National Health Insurance (NHIF).

### Ethical considerations

The analysed data in this study is subject to the following licenses/restrictions: The data contains potentially identifying and sensitive patient information. This has also been stipulated by the Local Institutional Review Board of KCMC hospital and the National Ethics Committee in Norway when establishing this birth registry. The College Research Ethics Review Committee of Kilimanjaro Christian Medical University College number 2606 and the Research Ethics Committees in Tanzania, ethical approval number NIMR/HQ/R.8a/Vo1. IX/4075. Verbal consent was sought from each mother prior to the interview, which was conducted just after the woman had given birth. Participation was voluntary and had no impact on the management women would receive. Mothers were free to refuse to reply to any single question.

### Data availability

Researchers can apply to access data from the KCMC Birth Registry data by contacting the Executive Director of the KCMC hospital in Tanzania, followed by submission of a proposal. The data are not publicly available due to privacy/ethical restrictions and are only available upon reasonable request.

### Collaboration

Birth registration is integral to society, and this is reflected in Sustainable Development Goal 16.9 which aims to provide legal identity for all, by the year 2030. From a societal and research perspective, the hospital statistics made available by the KCMC MBR become an important tool for monitoring progress toward achieving the Sustainable Development Goals (SDG) related to maternal and child health.

Data harmonization within and across countries is fundamental to researchers, and the interoperability of such data facilitates researchers to identify trends and patterns in core perinatal health outcomes, and compare such across different populations, and regions. The KCMC MBR has facilitated collaboration between countries within sub-Saharan Africa, the EU, and the UK and further afield, and is accessible to researchers for continued research. See

Appendix A for access to the full list of variables and Appendix B for the KCMC Medical Birth Register standardised questionnaire.

### Future plans for the KCMC MBR

1) Enhancements to birth registries are paramount for continued monitoring and forecasting of maternal and perinatal health care. Systematic improvements in the processes involved in recording and maintaining the data prioritising the linkage between MS Office access data with electronic data leads to improved data accuracy, quality, and efficiency.
2) Future studies and RCTs
3) Potentially collecting additional data on biomarkers such as adverse pregnancy outcomes like pre-eclampsia, and gestational diabetes will assist healthcare providers in developing individual patient care pathways and ultimately will improve patient outcomes.
4) The data has the potential to influence multigenerational studies and identify individual patient predispositions to adverse pregnancy outcomes based on familial history.

Ultimately, modernizing registration procedures can lead to more efficient data collection, entry, and retrieval processes. At present, information communication technology (ICT) product developers in Tanzania are working with clinicians and healthcare providers, trialling a mobile birth registry application. The ULTRA application is implemented through a specific configuration of the District Health Information System 2 (DHIS2) software. The benefit of an open-source delivery register functioning from a fully electronic input is to give institutions the freedom to register birth details from home settings through to tertiary care units, with a tablet, mobile phone, laptop, or desktop computer depending on the resources available to the healthcare worker - with or without access to the internet. The data is collected in aggregate form and anonymised for clinical and research purposes and uses. Implementing real-time or near-real-time reporting mechanisms can save time, improve efficiency and ensure birth information is promptly recorded and updated in the registry.

The overarching aim is to offer a solution to low-middle-income countries to further develop an interoperable and open-source digital healthcare system to enable other countries to adapt and replace paper-based management systems, reduce the workload and burden placed on healthcare providers for data collection and recording with monthly aggregated paper-based reports, to facilitate data harmonization within and across countries to monitor core perinatal health outcomes.

## Author’s contributions

ASK, BTM, IM, SW and BM designed the study. DB, MM, AS and CT drafted the manuscript. ASK, MM and MJM performed the statistical analysis. GM, RM, AKD, RTL, GN, AL, FMP and LM co-authors participated in editing the manuscript and approved the final manuscript for submission.

## Acknowledgments

We would like to express our gratitude to the midwives who contributed to the data collection process, as well as the women and children whose information enabled the availability of data used in this study. The authors also thank the staff at KCMC who captured data for the Medical Birth Registry in the electronic system.

We wish to further extend our thanks to the Centre for International Health and Public Health and Primary Health Care Department at the University of Bergen in Norway and the Department of Obstetrics and Gynaecology of the KCMC hospital in Tanzania for their collaboration to establish the KCMC medical birth registry, which has facilitated the availability of data to conduct this study.

## Appendix A: Subset of variables from the data dictionary

## Appendix B: KCMC Medical Birth Registry Questionnaire

## Appendix C

**PubMed List of Papers (Double click the table below, then click enable in order to view the list of Publication) PubMed Search:** “*medical birth reg*^*\**^*” OR “birth reg*^*\**^*” AND (Kilimanjaro OR Tanzania OR KCMC)* N=43 (2007 to 2024)

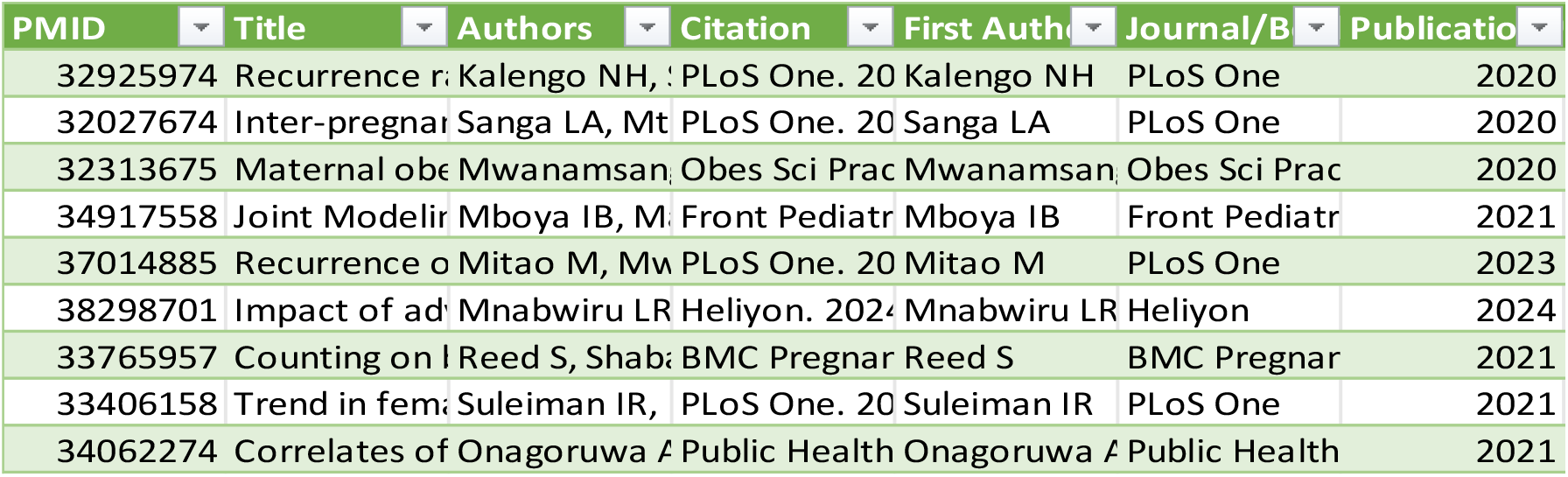

